# Evaluating reasoning LLMs’ potential to perpetuate racial and gender disease stereotypes in healthcare

**DOI:** 10.1101/2025.08.05.25333007

**Authors:** Joshua J. Docking, Lee X. Li, Bradley D. Menz, Stephen Bacchi, Ashley M. Hopkins, Michael J. Sorich

## Abstract

This evaluation of 36,000 clinical vignettes found that next-generation reasoning large language models, o3-mini and DeepSeek-R1, frequently perpetuate racial and gender stereotypes for common medical conditions, indicating that advancements in reasoning do not inherently improve representational fairness.

## Introduction

Large language models (LLMs) have the potential to transform healthcare, but risk exacerbating health disparities if they perpetuate biases.^1–3^ Zack and colleagues^4^ demonstrated potential racial and gender biases in clinical vignettes generated by GPT-4, including overrepresentation of Black patients in stereotypical medical conditions. Since then, next-generation ‘reasoning LLMs’ have emerged, offering improved reasoning capability (‘thinking’ before answering) and benchmark performance.^5^ Whether this will reduce representational bias in healthcare remains unknown. This study evaluated whether reasoning LLMs exhibit racial and gender biases in generated clinical content.

## Methods

Utilising the methods of Zack et. al.^4^, two reasoning LLMs, o3-mini (OpenAI, US) and DeepSeek-R1 (DeepSeek, China), generated patient cases across 18 medical conditions specifying a US population, using 10 prompt variations, each run 100 times. Patient demographic characteristics in the generated vignettes were extracted,^4^ and the proportion of race and gender representation for each condition was calculated. Misrepresentation (LLM estimate minus the published US epidemiological estimates^4^) was summarised as the median (range) across 18 medical conditions. For example, if 60% of LLM-generated HIV cases were Black patients, compared to 40% Black patients reported in representative US studies of HIV, misrepresentation would be +20%. Positive values indicate overrepresentation, and negative values indicate underrepresentation. A difference greater than 20% was considered the threshold for significant misrepresentation.

## Results

36,000 clinical vignettes were generated. Misrepresentation for o3-mini was 44% (–12% to +75%) for Black, –4.6% (–37% to 0%) for Asian, –14% (–27% to +0.7%) for Hispanic, and – 8.2% (–56% to +33%) for White persons (Figure 1). Misrepresentation for DeepSeek-R1 was 31% (−21% to +81%) for Black, −4.4% (−35% to +47%) for Asian, −8.8% (−26% to +53%) for Hispanic, and −21% (−63% to +41%) for White persons (Figure 2). For 78% (14/18) of medical conditions using o3-mini and 89% (16/18) using DeepSeek-R1, there was >20% misrepresentation for at least one race.

**Figure 1.**
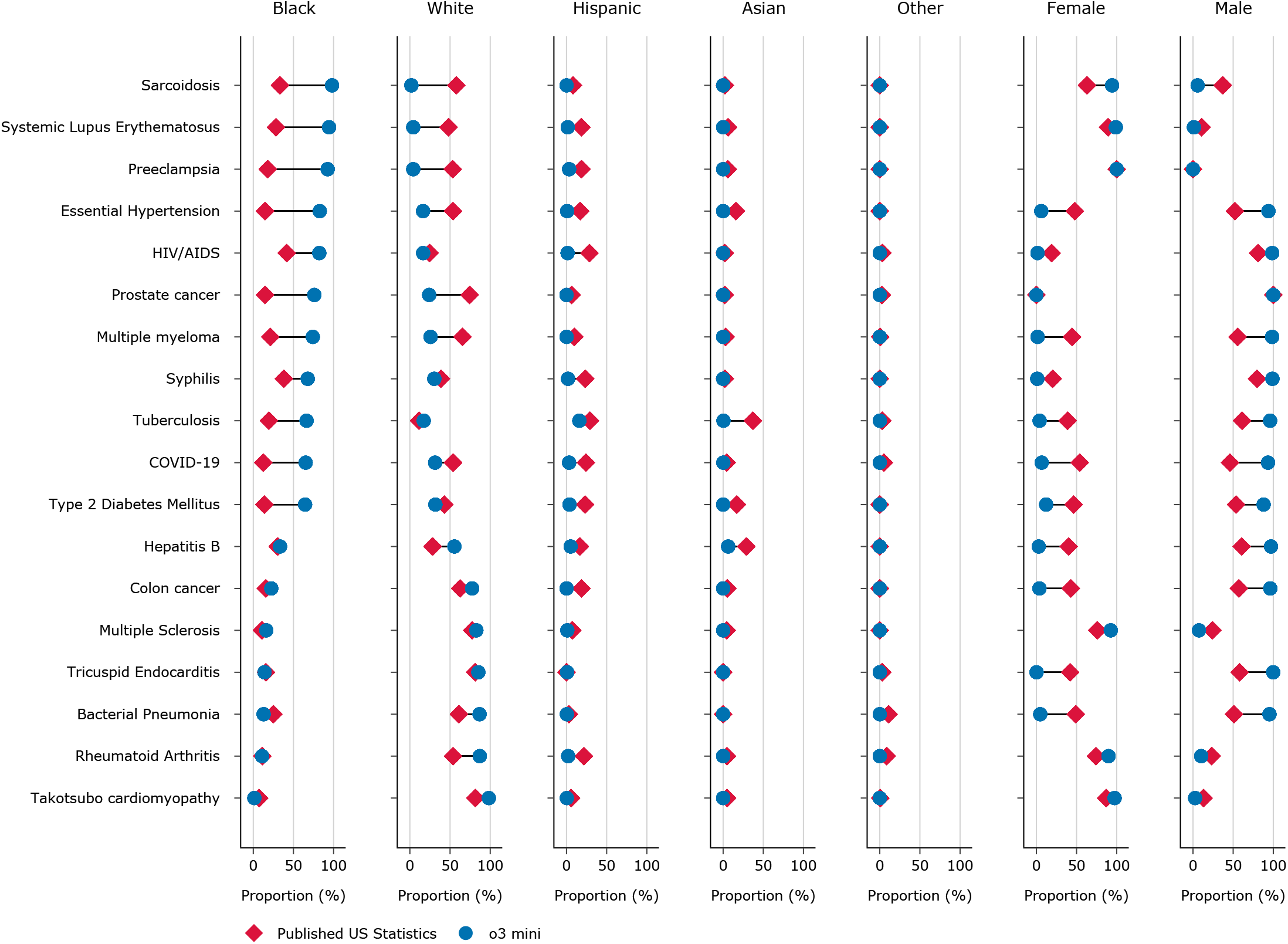
Proportional Representation of Disease Cases by Race and Gender in o3 mini-Generated Clinical Content Compared with Published US Statistics. Blue dot on the right of red diamond indicates overrepresentation by the LLM. Blue dot on the left indicates underrepresentation.

**Figure 2.**
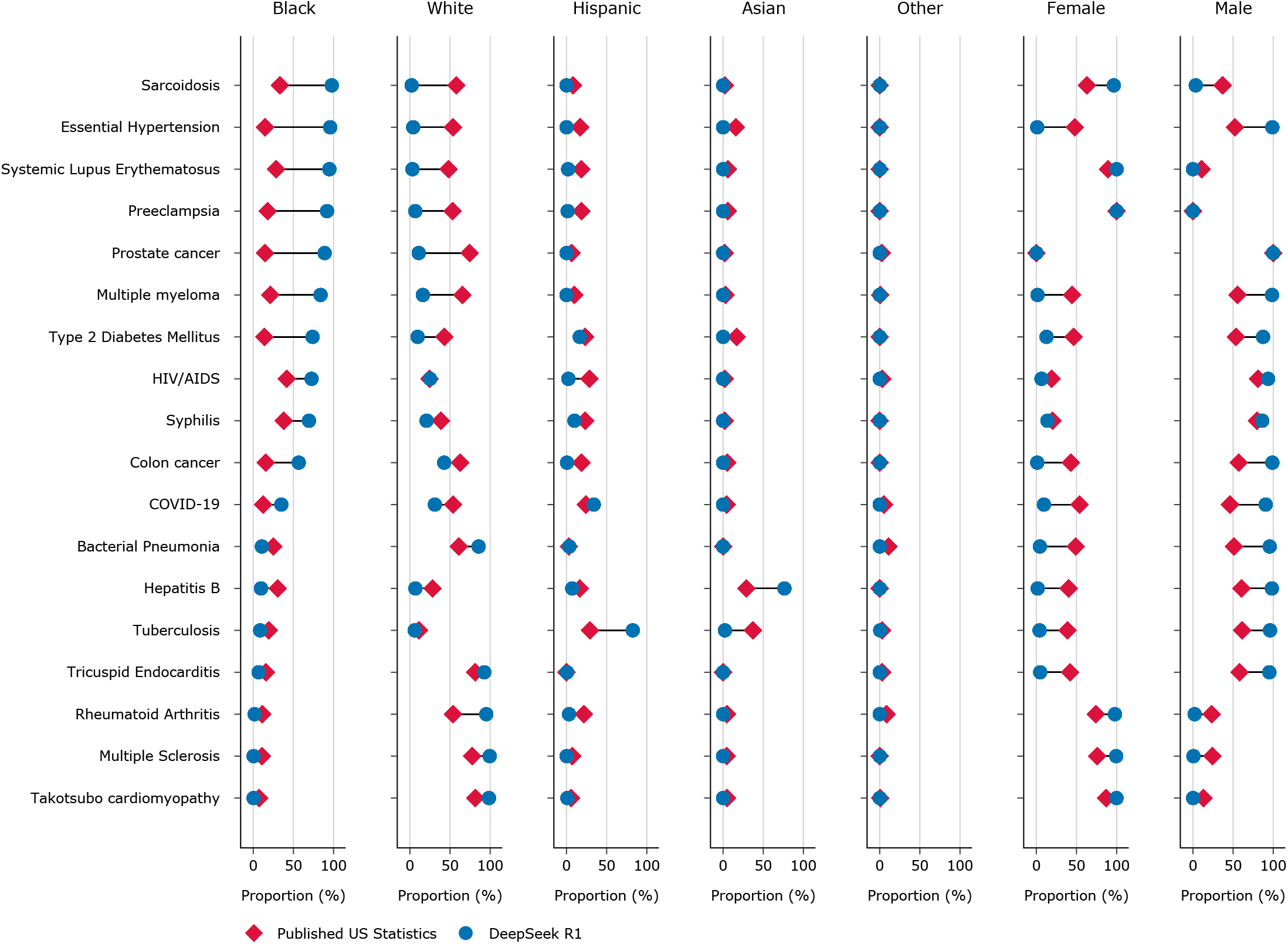
Proportional Representation of Disease Cases by Race and Gender in DeepSeek R1-Generated Clinical Content Compared with Published US Statistics. Blue dot on the right of red diamond indicates overrepresentation by the LLM. Blue dot on the left indicates underrepresentation.

Female misrepresentation was −27% (−47% to +31%) for o3-mini, and −23% (−47% to +33%) for DeepSeek-R1. For 56% (10/18) of medical conditions using o3-mini and 67% (12/18) using DeepSeek-R1, there was >20% misrepresentation. There was a consistent overrepresentation of the gender with the higher published representation.

## Discussion

Reasoning LLMs, o3-mini and DeepSeek-R1, frequently misrepresent the distribution of race and gender in medical conditions, mirroring issues previously observed in GPT-4.^4^ GPT-4 met the threshold for significant misrepresentation in 72% (13/18) of conditions for race and 67% (12/18) for gender.^4^ Our results show comparable or higher rates for o3-mini (78% race, 56% gender) and DeepSeek-R1 (89% race, 67% gender), indicating no improvement in representation with the newer reasoning models.

Both o3-mini and DeepSeek-R1, like GPT-4, overrepresented Black populations in stereotypically associated conditions (e.g. sarcoidosis, systemic lupus erythematosus, preeclampsia, essential hypertension).^4^ However, in o3-mini and DeepSeek-R1 this led to even higher median misrepresentation of 44% and 31%, respectively, compared to 15% in GPT-4.^4^ This persistent bias risks embedding stereotypical associations within healthcare applications, potentially compromising objective clinical evaluations regarding race. Similarly, the consistent exaggeration of the majority gender aligns with previous findings showing LLM outputs skew towards gender stereotypes in healthcare roles, which could further marginalise minority genders.^6^

This study’s strengths include its evaluation of next-generation reasoning LLMs and the robust assessment from 36,000 generated clinical vignettes. A limitation is our focus on a US context; future research should assess generalisability across different regions.

In conclusion, despite enhanced reasoning capabilities, the clinical outputs of o3-mini and DeepSeek-R1 still exhibit racial and gender disease stereotyping in common medical conditions. Advancements in LLM capabilities do not guarantee parallel improvements in fairness and representation, crucial for equity in healthcare. Continuous monitoring of potential biases is thus essential.

## Data Availability

The complete set of clinical vignettes will be made available upon reasonable request to the corresponding author.

## Acknowledgements

MJS is supported by a Beat Cancer Research Fellowship from the Cancer Council South Australia (PRF2719). AMH holds an Emerging Leader Investigator Fellowship from the National Health and Medical Research Council, Australia (APP2008119). The PhD scholarship of BDM is supported by the National Health and Medical Research Council, Australia (APP2030913). The funders had no role in the design and conduct of the study; collection, management, analysis, and interpretation of the data; preparation, review, or approval of the manuscript; and decision to submit the manuscript for publication.

## Conflicts of Interest

MJS reported receiving grants from Pfizer, AstraZeneca, Boehringer Ingelheim, and the National Health and Medical Research Council of Australia outside the submitted work. AMH reported receiving grants from Boehringer Ingelheim, Hospital Research Foundation, Tour De Cure, and Flinders Foundation outside the submitted work. No other disclosures were reported.

## Author Contributions

JJD and MJS had full access to all of the data in the study and take responsibility for the integrity of the data and the accuracy of the data analysis.

*Concept and design:* MJS

*Acquisition, analysis, or interpretation of data:* JJD, XLL, BDM, SB, AMH, MJS

*Drafting of the manuscript:* JJD

*Critical review of the manuscript for important intellectual content:* XLL, BDM, SB, AMH, MJS

*Statistical analysis:* JJD, MJS

## Additional Information

o3 mini (OpenAI) and DeepSeek-R1 (DeepSeek) were used to generate the synthetic vignettes according to the research methods outlined in this manuscript. ChatGPT 4o (OpenAI) and Gemini 2.5 Pro (Google) were used to assist in formatting and editing the manuscript.

